# Research Letter: Therapeutic targets for haemorrhoidal disease: proteome-wide Mendelian randomisation and colocalization analyses

**DOI:** 10.1101/2023.06.19.23291373

**Authors:** Shifang Li, Meijiao Gong

**Affiliations:** Laboratory of Immunology and Vaccinology, FARAH, ULiège, Liège 4000, Belgium

**Keywords:** Haemorrhoidal disease, Mendelian randomisation, ERLEC1, myxoedema

## Abstract

Human haemorrhoidal disease (HEM) is a common anorectal pathology. However, the etiology of HEM, as well as its molecular mechanism, remains largely unclear. In this study, we applied a two-sample bi-direction Mendelian randomisation (MR) analysis to estimate the causal effects of 4907 plasma proteins on HEM outcomes and investigated the mediating impacts of plasma proteins on HEM risk factors to uncover potential HEM treatment targets by integrating GWASs statistics of HEM and plasma protein levels. Following MR analysis, our study identified 5 probable causal proteins associated with HEM. ERLEC1 and ASPN levels were genetically predicted to be positively and inversely associated with HEM risk, respectively, with strong evidence of colocalization (H4>0.9). The findings of an independent cohort corroborate the causal relationship between these two proteins and HEM. Furthermore, gene expression analysis of haemorrhoidal tissue and normal specimens revealed that ERLEC1 but not ASPN were differentially expressed. By analyzing single-cell ERLEC1 expression in human rectum tissues, ERLEC1 was found to be highly expressed in transient-amplifying cells. Interestingly, a genetically greater risk of myxoedema was linked to an elevated risk of HEM. However, there was no evidence that dorsalgia, hernia, diverticular disease, and ankylosing spondylitis were causally associated with HEM. Furthermore, no association was found between myxoedema and the genetically predicted ERLEC1 and ASPN levels. Overall, this study identified some causal associations of circulating proteins and risk factors with HEM by integrating the largest-to-date plasma proteome and GWASs of HEM. The findings could provide further insight into understanding biological mechanisms for HEM.

---

Human haemorrhoidal disease (HEM) is a common anorectal disorder. Recently, Zhang *et al*. reported the first and largest genome-wide association study (GWAS) with haemorrhoidal disease (HEM), and these data offered us a resource for understanding the genetic risk factors for HEM.^1^ However, the etiology of HEM, as well as its molecular mechanism, remains primarily unclear.^2^ In addition, the identification of genes with therapeutic effects needs to be conducted. In recent years, by incorporating protein quantitative trait loci (pQTLs) into MR analysis, such an approach has been successfully used to prioritize therapy targets.^3,4^ Here, using a two-sample bidirectional Mendelian randomisation (MR) analysis, we estimated the causal effects of 4907 plasma proteins on HEM outcomes, and investigated the effects of plasma proteins that may mediate the impact of risk factors on HEM in order to identify potential therapeutic targets for HEM.

As stated in the **supplementary methods**, 4907 proteins (*cis*-pQTLs) were used as instrumental variables for exposure and HEM as the outcome to estimate the causal effect of plasma protein levels on HEM in a proteome-wide context using MR analysis.^5-8^ Our study revealed 5 potential causative proteins at the Bonferroni-corrected threshold of *p*<1.01×10^−5^, including 3 negative and 2 positive associations (**figure 1A-1B**). MR analysis, for example, revealed that genetically predicted ERLEC1 levels were linked to an increased risk of HEM (*p*=5.18e-07). To determine whether the identified relationships of the circulating protein with HEM shared causative variations, colocalization analysis was carried out and a high level of support for colocalization evidence was discovered between two proteins (ERLEC1 and ASPN) and HEM (H4>0.9) (**figure 1C**). The findings of the INTERVAL cohort corroborated the causal relationship between these two proteins and HEM (**figure 1D**).^9^ Interestingly, the deCODE study’s lead cis-pQTL for the ERLEC1 (rs2542580) but not ASPN (rs10992273) were not found to be associated with all available secondary traits (**supplementary table1**). Gene expression analysis of haemorrhoidal tissue and normal specimens revealed that ERLEC1 but not ASPN were differentially expressed after controlling for gender and BMI (**figure 1E**), further supporting that a high ERLEC1 expression level was associated with an increased risk of HEM. Following that, we investigated the tissues in which ERLEC1 is expressed in bulk tissues using GTEx v8 (https://gtexportal.org/), and found that ERLEC1 was considerably expressed in multiple tissues, including the small intestine and colon, as compared to the whole blood (*p*<0.001) (**figure 1F**). To further understand the origin of ERLEC1, single-cell ERLEC1 expression was assessed in human rectum tissues, and ERLEC1 was found to be highly expressed in transient-amplifying (TA) cells (*p*< 0.05) (**figure 1G**).^10^

**Figure 1.**
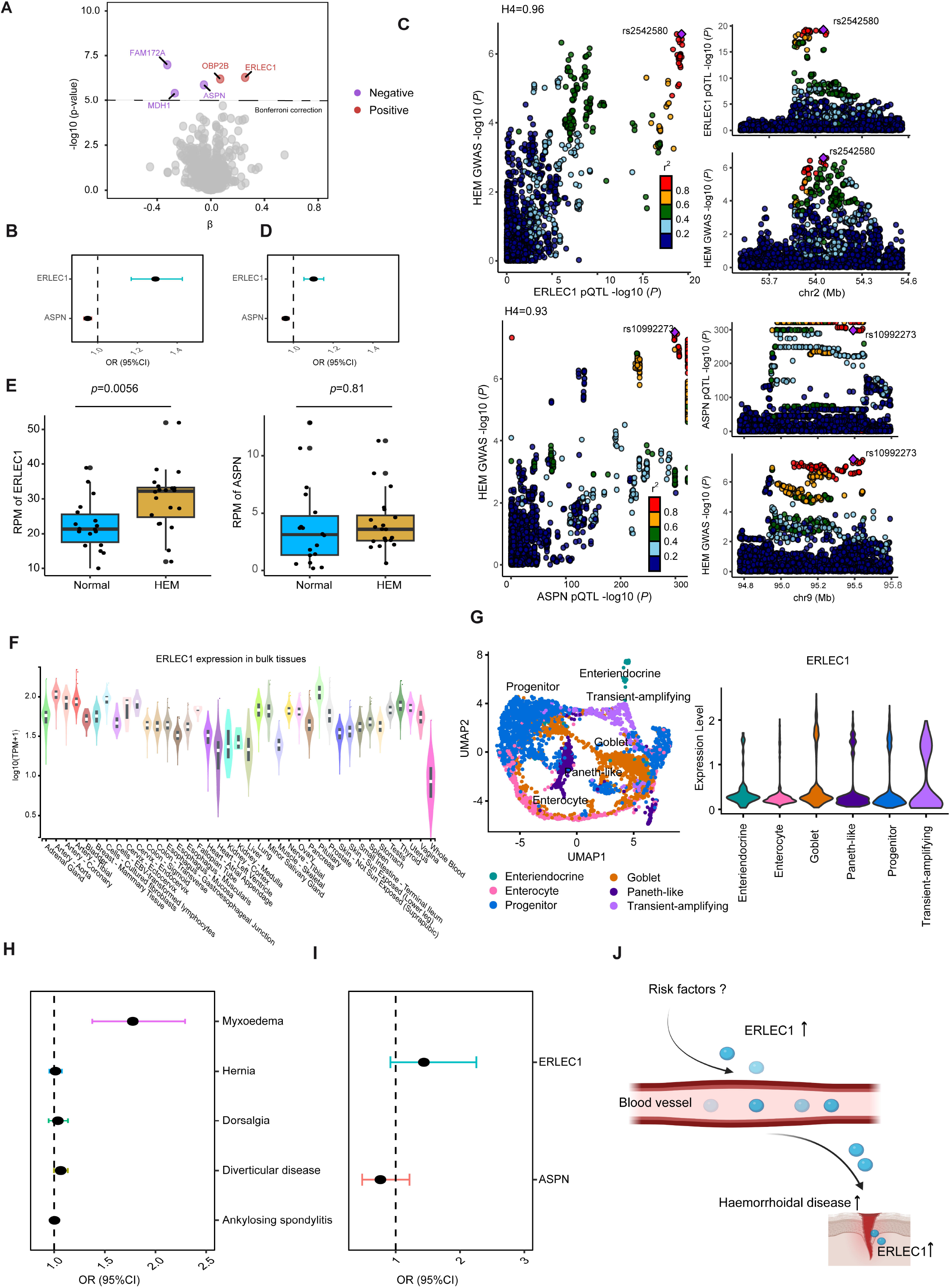
Mendelian randomisation results. (**A**) The effect of plasma protein levels on HEM. Volcano plot indicating the effect of plasma protein on HEM using MR analysis. (**B**) Forest plots shows the effect of plasma ERLEC1 and ASPN levels on HEM. (**C**) Colocalization analysis of ERLEC1 levels (Up) and ASPN (Down). (**D**) Forest plots shows the effect of plasma ERLEC1 and ASPN levels on HEM using INTERVAL cohort. (**E**) Boxplot shows differentially expressed genes in HEM patients when compared to healthy individuals. *p*-values were corrected the effect of gender and BMI using linear model. (**F**) The violin plot depicts ERLEC1 gene expression across multiple bulk tissues. (**G**) Data visualization of cell populations in human rectum tissues using UMAP (left) and gene expression of ERLEC1 in different cell types (right). (**H**) Forest plots showing the causal effect of chosen risk factors on HEM. (**I**) Forest plots for the effect of myxoedema on plasma ERLEC1 and ASPN levels. (**J**) Schematic illustration of the proposed model in the study. HEM, haemorrhoidal disease.

In order to investigate whether the causal protein mediates the effect of risk factors on HEM, the causal risk factors for HEM were first identified. 5 clinical traits that genetically correlated with HEM were selected (**supplementary methods**), with instrumental variables generated from GWASs confined to European populations. It was discovered that a genetically greater risk of myxoedema was linked to an elevated risk of HEM (*p*<0.05) (**figure 1H**). Although genetic correlations with HEM were reported,^1^ there was no evidence that dorsalgia, hernia, diverticular disease, and ankylosing spondylitis were causally associated (*p*>0.05). In order to identify the protein related to HEM risk factors, we conducted MR analysis again on 2 plasma proteins impacting HEM with myxoedema. After filtering, there was a lack of evidence that myxoedema had a causal relationship with these two plasma proteins (**figure 1I**).

Overall, by integrating the largest-to-date plasma proteome and GWAS of HEM, we discovered that ERLEC1 could serve as prospective protein therapeutic targets for HEM. In-depth research is needed to investigate the mechanisms by which putative risk factors affect HEM (**figure 1J**).

## Supporting information

The significant MR summary statistics obtained in this study.

## Data Availability

All data produced in the present work are contained in the manuscript.

## Contributors

SF was involved in conceptualization. SF and MJ were involved in the formal analysis. SF was involved in writing, reviewing, and editing.

## Acknowledgments

The authors would like to thank all of the researchers who contributed to the GWAS datasets used in this study for making them available for research purposes.

## Supplementary Methods

The statistics method used in the study.

**Supplementary Tables1** The significant MR summary statistics obtained in this study.

## Supplementary Methods

### GWASs of haemorrhoidal disease and risk factors

We used recently published large-scale genome-wide associations (GWASs) for haemorrhoidal disease (HEM).^1^ This GWAS summary statistics were derived from 944,133 European ancestry individuals (Ncase = 218,920 and Ncontrol = 725,213) from 5 cohorts and downloaded from the GWAS Catalog (https://www.ebi.ac.uk/gwas/, access ID: EFO_0009552). Diverticular disease of the intestine, ankylosing spondylitis (AS), dorsalgia, hernia, and myxoedema were evaluated as potential causal risk factors associated with HEM in order to determine the probable causal risk factors. All GWASs for the five risk factors were obtained from the ieu open gwas project (https://gwas.mrcieu.ac.uk/datasets/). The summary statistics of the large GWAS (14,357 cases and 182,423 controls) were used for diverticular disease of the intestine (access ID: finn-b-K11_DIVERTIC). The GWAS for ankylosing spondylitis (access ID: finn-b-M13_ANKYLOSPON) have a sample size of 1,462 cases and 164,682 controls. The GWAS for myxoedema (access ID: ieu-b-4877) has a sample size of 311,629 cases and 321,173 controls. The GWAS for dorsalgia (access ID: finn-b-M13_DORSALGIA) included 193467 individuals, with 28,785 cases and 164,682 controls. A total of 218792 individuals were reported with GWAS of hernia (access ID: finn-b-K11_HERNIA), including 28,235 cases and 190,557 controls.

### Plasma protein quantitative trait loci (pQTL) data

To conduct proteome-wide Mendelian randomisation (MR), we first obtained genetic instrumental variables using the protein quantitative trait loci (pQTL) data generated by Ferkingstad *et al*.^2^ The largest-to-date pQTL analysis on plasma proteome (a total of 4907 proteins) in 35,559 Icelanders was performed in their study, and an amount of 18,084 pQTL associations between genetic variation and protein levels in plasma were identified. A total of 4907 pQTLs were successfully downloaded from the deCODE study using aria2c.^3^ To minimize the risk of horizontal pleiotropy, instrumental variables to *cis*-pQTLs (SNPs located within a 500 kb window from the target gene body) of protein were selected for the following analysis. In order to validate the MR results for ASPN and ERLEC1 using independent study, the *cis*-pQTLs of ASPN and ERLEC1 were obtained from the INTERVAL cohort and used for the following analysis.^4^

### Mendelian randomisation analysis

MR analysis is an analytical method that uses genetic variation as an instrumental variable (IV) to estimate causal effects. It overcomes the limitations of measurement error and confounding factors that are common in observational studies and is widely used to assess causal relationships.^5^ In this study, the TwoSampleMR package (v0.5.6, https://mrcieu.github.io/TwoSampleMR/) was used for MR analysis.^6^ The instrumental variables that determined the exposure in each MR study were specified as genome-wide significant (*p* ≤ 5e-08) SNPs. SNPs in the human major histocompatibility complex (MHC) region at chromosome 6: 28,477,797-33,448,354 (GRCh37) were excluded from the analysis due to its complex linkage disequilibrium (LD) structure. Using the 1000 Genomes Project European reference panel and an LD threshold of r^2^ <0.001 with a clumping window of 10,000 kb, PLINK v.1.9 (http://pngu.mgh.harvard.edu/purcell/plink/) was employed to derive instrumental variables.^7-8^ F-statistics were used to determine the strength of each SNP’s association with exposure, and F-statistics of more than 10 were considered strong. For the main MR analysis, the inverse variance weighted approach for proteins with two or more instrumental variables and the wald ratio method for proteins with a single instrumental variable was used for evaluating the causal influence of exposure on outcome. In addition, in the case of more instrumental variables used in MR analysis, four additional MR methods (weighted median, simple mode, weighted mode, and MR-Egger method) were used to assess the reliability of the primary results. For exposures with multiple IVs, we additionally investigated heterogeneity across variant-level MR estimations with the “mr_heterogeneity()” function in the TwoSampleMR package (Cochrane’s Q test). In addition, a pleiotropy test was performed using MR Egger analysis to determine whether there is horizontal pleiotropy among IVs. Meanwhile, “phenoscanner” (https://github.com/phenoscanner/phenoscanner) was to be utilized to determine any pleiotropy of SNPs used in the MR analysis. An SNP was regarded to be pleiotropic when the reported SNP-traits association was genome-wide significant (*p*≤ 5e-08) in the European population.

Finally, in the event there were more than two IVs in exposure, a leave-one-out analysis was performed, and the MR findings of the remaining IVs were calculated by deleting the IVs one by one to ensure the robustness of the MR data. To acquire robust evidence for the casual estimation, MR findings that meet all of the following criteria were chosen as described by Yoshiji and others: (1) no pleiotropy was found using MR-Egger regression (*p*>0.05); (2) results with an I^2^ < 50% (no substantial heterogeneity); (3) leave-one-out analysis MR *p<*0.05 after removing outliers; and (4) reverse MR *p>*0.05.^9^ The same procedure as mentioned above was utilized to explore the causal effect of the given exposure and associated outcome in the reverse MR analysis. *p*-values less than a Bonferroni adjusting (*p*=1.01×10^−5^ (0.05/4,907)) are deemed significant for multiple testing.

### Colocalization analysis

The coloc R package was employed to investigate whether the reported relationships between proteins and HEM were driven by linkage disequilibrium.^10^ The analysis offers posterior probability for each hypothesis tested: no association in either group (H0), pQTL only (H1), the GWAS of HEM only (H2), associations with both GWAS but by separate causal signals (H3), and associations with both GWAS but by the same signals (H4).^11^ A higher H4 (H4>0.8) was considered as strong evidence for colocalization, implying a shared variation between the two phenotypes.^10,11^

### Differentially expressed genes analysis in bulk tissues

The GSE154650 dataset was downloaded from NCBI Gene Expression Omnibus (GEO) and analyzed using the R program.^12^ The RPM value of ERLEC1 and ASPN were further subjected to linear model analysis to investigate the differential gene expression in HEM and healthy individuals after correcting for the effects of gender and BMI. The expression data of ERLEC1 from 39 tissues across 838 individuals were obtained from the GTEx v8 (https://gtexportal.org/).^13^ Mann-Whitney U test was performed to determine the significance of ERLEC1 expression differences between the two groups, and *p<*0.01 was declared significant.

### scRNA-sequencing analysis of human rectum tissues

For processing scRNA data (GSE125970), the raw data of the gene expression matrix was first downloaded from NCBI Gene Expression Omnibus (GEO) and converted into a Seurat object using the R Seurat package.^14,15^ Low-quality cells were eliminated if they met any of the following requirements: (1) 3000 UMIs; (2) 200 genes; and (3) >50% of UMIs derived from the mitochondrial genome. UMI counts were normalized using the NormalizeData function, and the top 2000 features with the greatest cell-to-cell variation were calculated using the FindVariableFeatures function. To correct the batch effects among samples, the “FindIntegrationAnchors” and “IntegrateData” functions were employed. Following that, the ScaleData function was used to scale and center features in the datasets, and the RunPCA function with default parameters was used to reduce dimensionality. The data were then used for nonlinear dimensional reduction with the RunUMAP function and cluster analysis with the FindNeighbors and FindClusters functions. The FindAllMarkers function was used to identify differentially expressed genes (DEG) for a given cluster. The clusters were labeled in the same way that Wang *et al*. did in their study.^15^

